# Vitamin D levels, assessment of evidence for an association between vitamin D deficiency and atherosclerosis: a two-way Mendelian randomization study

**DOI:** 10.1101/2025.03.19.25324287

**Authors:** Chunhui Liu, Xvpeng Huang, Hongguang Jin, Yongsheng Huang

## Abstract

**Background and Aim:** Previous studies have shown an association between vitamin D and atherosclerosis, but whether there is a causal relationship between the two is not yet clear and requires further study.The analytical approach of a two-sample Mendelian randomization study was used to further investigate the effect of vitamin D on AS.

**Methods and Results:** Vitamin D levels, vitamin D deficiency, and AS (including coronary atherosclerosis, cerebral atherosclerosis, aortic atherosclerosis, and peripheral atherosclerosis) were analyzed in samples from the Gwas database, all of which were of European origin. To assess the presence of reverse causality, data from the vitamin D-related samples were analyzed by reverse MR as an outcome.

Forward MR analyses of vitamin D levels, vitamin D deficiency, and atherosclerosis were negative, i.e., there was no positive causal association. For sensitivity analysis, Cochran’s Q test suggested no heterogeneity, and MR-Egger showed no evidence of horizontal pleiotropy. In contrast, our subsequent inverse MR analysis showed a causal association between peripheral atherosclerosis and vitamin D deficiency, i.e., peripheral atherosclerosis can cause the development of vitamin D deficiency. Sensitivity analyses did not reveal the presence of heterogeneity and horizontal pleiotropy. Leave-one-out analysis showed good robustness of the corresponding SNPs without bias. MR analyses between the other three types of atherosclerosis and vitamin D levels, and vitamin D deficiency, resulted in no causal association.

**Conclusions:** Our MR analysis identified peripheral atherosclerosis as a factor in the pathogenesis of vitamin D deficiency, which provides a reference for subsequent research and treatment of this disease.

**Lay summary:** We found through MR means that there is no positive causality between vitamin D and atherosclerosis, and it is unexpected that it is found that the outer peripheral atherosclerosis can cause vitamin D deficiency.

## 1. Introduction

Atherosclerosis (AS) is a pathological change mainly characterized by the appearance of plaques in the arterial wall, causing narrowing or blockage of arterial blood vessels and thus triggering symptoms. Depending on the site of occurrence, it can be divided into coronary atherosclerosis, cerebral atherosclerosis, aortic atherosclerosis, and peripheral atherosclerosis. Modern research has demonstrated that the pathogenesis of AS involves a combination of inflammation and immune response [1]. Atherosclerosis-associated cardiovascular disease is the leading factor contributing to the global increase in mortality, with 460,000 related deaths globally over seven years, accounting for approximately one-third of all deaths, as reported by the World Health Organisation in 2016 [2]. In the United States, cardiovascular disease leads to mortality rate, and related surveys have shown that the prevalence of cardiovascular disease among inflammatory disease individuals in the United States has been high for nearly 20 years, from 1999 to 2018 [3]. Vitamin D is a vitamin that promotes calcium absorption in the body and undergoes 25-hydroxylation in the liver to 25(OH)D (osteoclastic diol) to participate in circulation. Metabolism in the kidneys can be converted to active 1,25(OH)2D (oestriol) through the process of 1-alpha-hydroxylation, a compound that acts by binding to the vitamin D receptor (VDR) in nucleated cells and can be found widely in cells of the brain, heart, and other organs [4].

Vitamin D is associated with anti-inflammatory, immunomodulatory, and antioxidant effects, so its associated deficiency may contribute to the development of AS. Various studies have shown a correlation between vitamin D levels and the incidence of cardiovascular events, making it particularly important to investigate its relationship with AS [5, 6]. Previous studies have found that vitamin D is inextricably linked to the development of AS, however, a causal relationship between the two has not been clearly established [7]. In this paper, we explore the causal relationship between vitamin D and AS based on Mendelian randomization (MR) study, which can help in clinical treatment and subsequent research.

## 2 Materials and Methods

### 2.1 Design Idea

A two-sample Mendelian randomization analysis was conducted using genome-wide (GWAS) data published on publicly available databases. The screened instrumental variables include the three assumptions of correlation, exclusivity, and independence. (i) the selected instrumental variables must be strongly correlated with the exposure factors; (ii) the instrumental variables can only affect the outcome through the exposure factors, and cannot have a direct effect alone; and (iii) the confounding factors between the exposure and the outcome cannot have an effect on the instrumental variables. The cases involved in the sample data in this study had signed authorized consent, ethical review, and approval. Figure 1 shows the flow chart of the study.

**Fig. 1.**
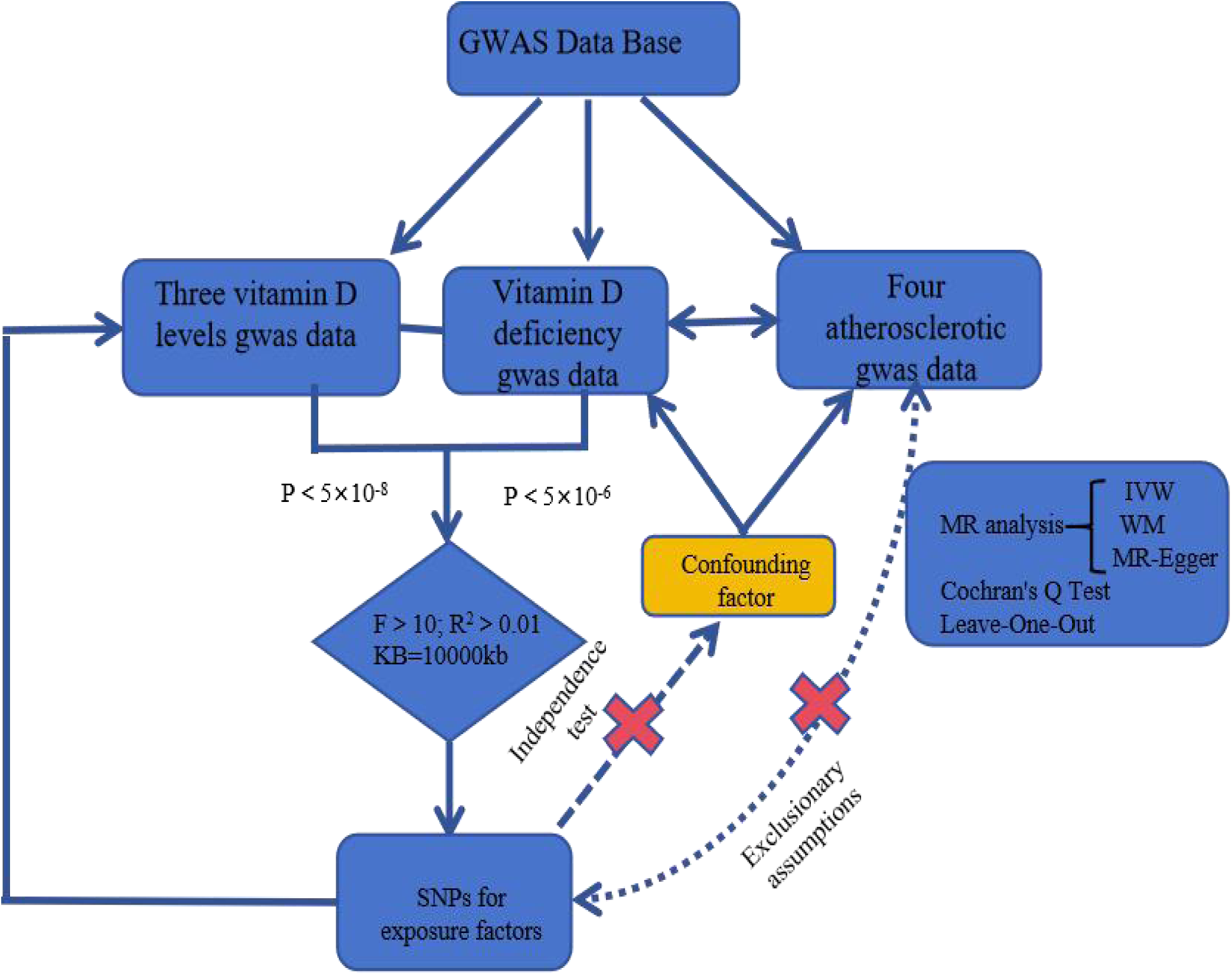
Flow chart of this study.

### 2.2 Data sources

#### 2.2.1 Vitamin D level GWAS data acquisition

Vitamin D levels were obtained from the GWAS public database for European populations. Vitamin D GWAS data were obtained from the IEU database (https://gwas.mrcieu.ac.uk/) for three different samples, including the corresponding immunofluorescence assay data for 25-hydroxyvitamin D (ID GCST90000618) containing 496,946 samples, and the Diasorin assay data (ID ieu-b-4812) containing 441,946 samples, 946 samples, Diasorin assay data (ID ieu-b-4812) containing 441,291 samples, and a data from meta-analysis (ID ebi-a-GCST005367) containing 79,366 samples [8, 9]. Vitamin D deficiency sample data were obtained from the FinnGen Biobank (ID finn-b-E4_VIT_D_DEF), available online at https://gwas.mrcieu.ac.uk/datasets/finn-b-E4_VIT_D_DEF/), containing 209,789 samples (182 cases, 209,607 controls) [10].

#### 2.2.2 Atherosclerosis GWAS data acquisition

Atherosclerosis-related sample data were obtained from the GWAS data (https://gwas.mrcieu.ac.uk/), with cases from populations of European origin. These included coronary atherosclerosis data (sample size 361,194), cerebral atherosclerosis data (sample size 218792), aortic atherosclerosis data (sample size 150765), and peripheral atherosclerosis data (sample size 168832).

### 2.3 Screening for genetic variants

(I) SNPs associated with vitamin D levels were screened with a p-value of <5×10-8, and SNPs associated with vitamin D deficiency were screened with a p<5×10-6 as a condition [11], and SNPs with significance were selected; (II) the linkage disequilibrium coefficient (LD) was set to r2=0.001, and the distance of the LDs was set to 10,000 kilobases to ensure the independence of the SNPs; (III) an F-value was calculated to assess the strength of the instrumental variable (IV) when performing MR analyses to determine the presence of weak variable bias. An F-value >10 indicates a strong association.

### 2.4 Statistical analysis methods

#### 2.4.1 MR and leave-one-out analysis

In this two-sample Mendelian randomization study, four MR analysis methods were used including inverse variance weighting (IVW), MR-Egger regression, weighted median, and weighted mode methods. The inverse variance weighting (IVW) method is the main method among MR, which further assesses the effect of exposure on outcome by calculating the Wald ratio for each SNP, which is combined with the inverse of the variance. The weighting model involves clustering SNPs with similar individual ratios into groups, calculating the cubic variance weighting of the SNPs in each group, and estimating the corresponding causality based on the weights of the SNPs. The leave-one-out method of analysis is to exclude SNPs that do not meet the requirements one by one, and for the remaining SNPs to analyze and tested with inverse variance weighting to assess the degree of influence of the SNPs on the overall effect, and if statistical insignificant differences are found, it means that the results of the corresponding MR analyses are stable.

#### 2.4.2 Sensitivity analysis

The Cochran’s Q test was used to assess whether there was heterogeneity between the samples for exposure and outcome. MR-Egger regression was used to fit a linear function by calculating the association between each SNP and the exposure and outcome, with the sloped distance representing the causal effect, and the MR-Egger intercept to determine whether there was a level of multinomial between the SNPs, if the corresponding P < 0.05, it indicates that there is horizontal pleiotropy between the two, i.e. the corresponding outcome occurs without the interference of the exposure factor.

### 2.5 Reverse Mendelian randomization study

To clarify the causal relationship between vitamin D levels, vitamin D deficiency, and atherosclerosis (coronary atherosclerosis, cerebral atherosclerosis, aortic atherosclerosis, and peripheral atherosclerosis), we carried out a reverse Mendelian randomization analysis by considering the four types of atherosclerosis as the exposure factors and vitamin D as the outcome.

## 3 Results

### 3.1 Screening of instrumental variables

For vitamin D level-related SNPs a p-value of <5×10-8 and for vitamin D deficiency-related SNPs a p<5×10-6 were screened to ensure an F-value of >10. Specific results are shown in Supplementary **Table 1**.

### 3.2 MR analysis

By MR analysis, the results of the present study did not show a causal relationship between vitamin D levels and atherosclerosis. The average F-statistic was >10, indicating that there was no effect of weak bias. The forest plots obtained by MR analysis are shown in **Figure 2**. The MR-Egger intercepts did not show evidence of horizontal pleiotropy, and the corresponding intercepts and sloped distances are shown in **Figure 3**. The results were analyzed by the IVW method, and the p-values were all greater than 0.05.The results of the data were analyzed in a weighted mode, and the results still did not suggest valid evidence of a causal relationship between vitamin D levels and atherosclerosis, and the specific information is shown in **Table 1**. The results of the data were analyzed in a weighted mode, and the specific information is shown in **Table 1**. The data were analyzed by the leave-one-out method to test for data of unstable nature. The robustness of the data between the corresponding SNPs for vitamin D deficiency and the four atherosclerosis data was good, whereas the data between the other three SNPs related to vitamin D levels and the four atherosclerosis data lacked robustness, and we removed the corresponding SNPs and repeated MR analyses still did not suggest any evidence of causality, and the results of the leave-one-out analyses are shown in **Figure 4**.

**Figure 2.**
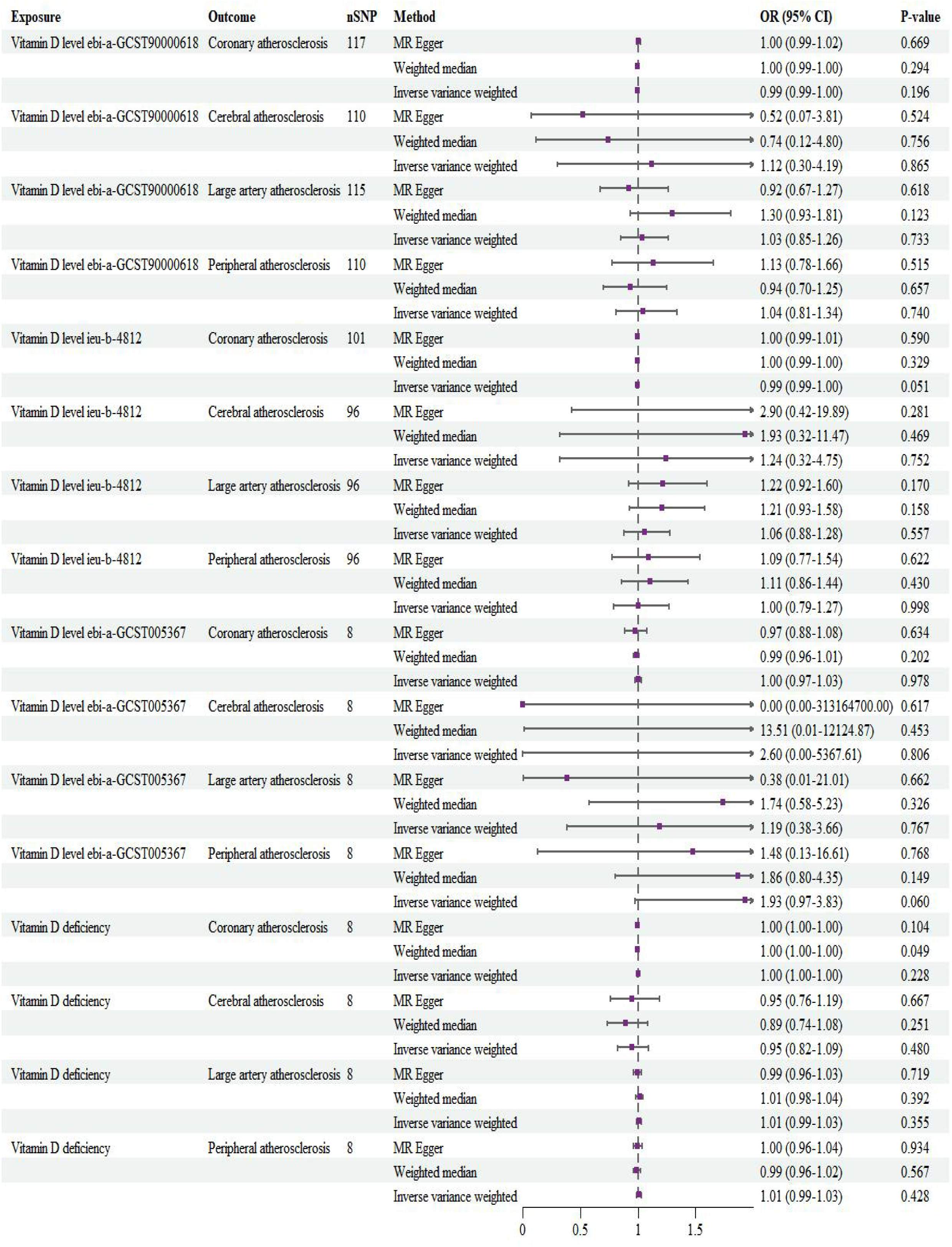
Forest map generated by MR analysis.

**Figure 3.**
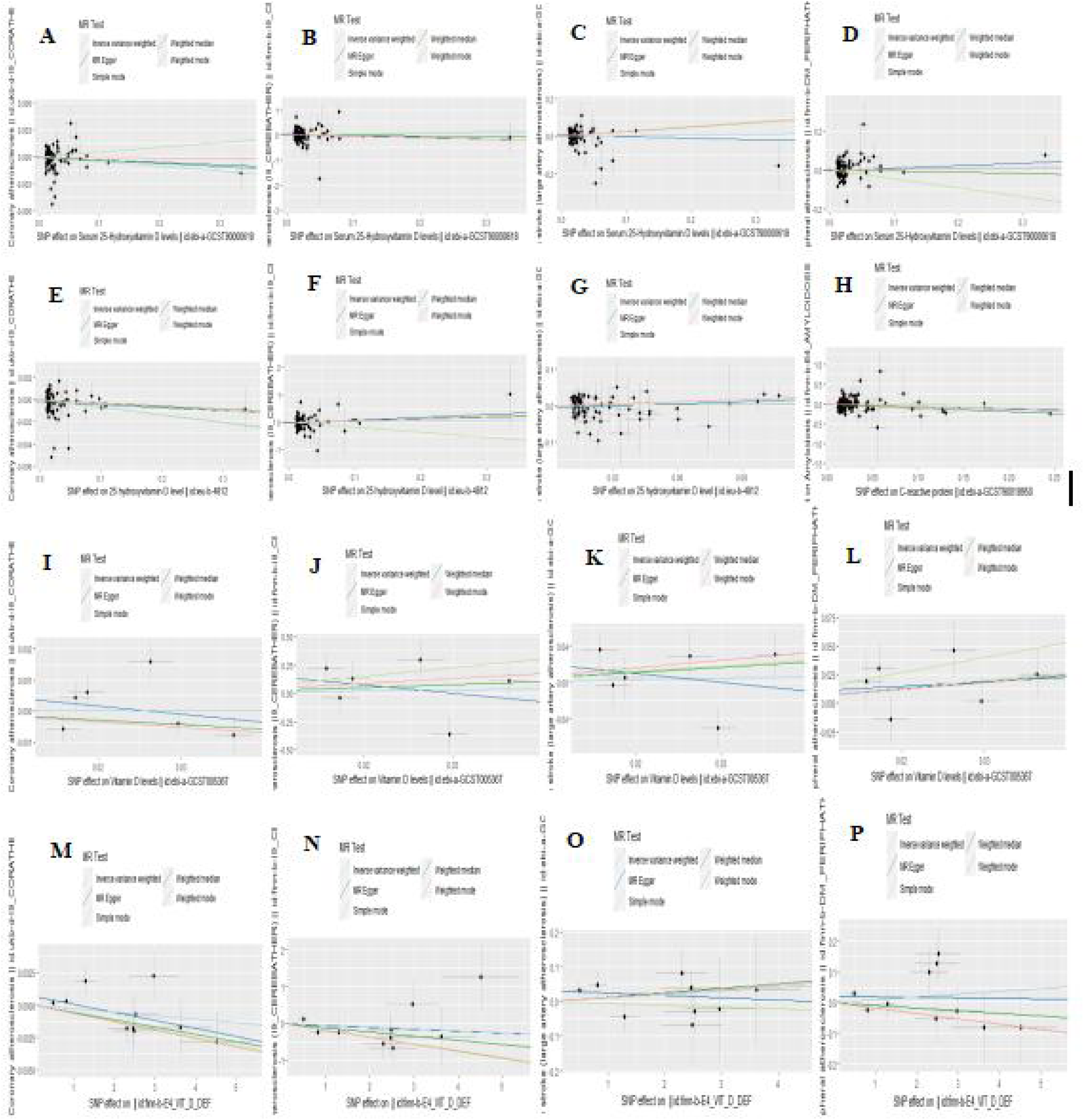
Scatter plot of vitamin D levels and MR analysis of atherosclerosis ABCD: ID ebi-a_GCST90000618: A. Coronary atherosclerosis, B. Cerebral atherosclerosis, C. Large artery atherosclerosis, D. Peripheral atherosclerosis; EFGH: ID ieu_b_4812: E. Coronary atherosclerosis, F. Cerebral atherosclerosis, G. Large artery atherosclerosis, H. Peripheral artery atherosclerosis; IJKL: ID ebi-a-GCST005367: I. Coronary artery atherosclerosis, J. Cerebral artery atherosclerosis, K. Large artery atherosclerosis, L. Peripheral artery atherosclerosis; MNOP: ID finn-b-E4_VIT_D_DEF: M. Coronary artery atherosclerosis, N. Cerebral artery atherosclerosis sclerosis, O. aortic atherosclerosis, P. peripheral atherosclerosis.

**Figure 4.**
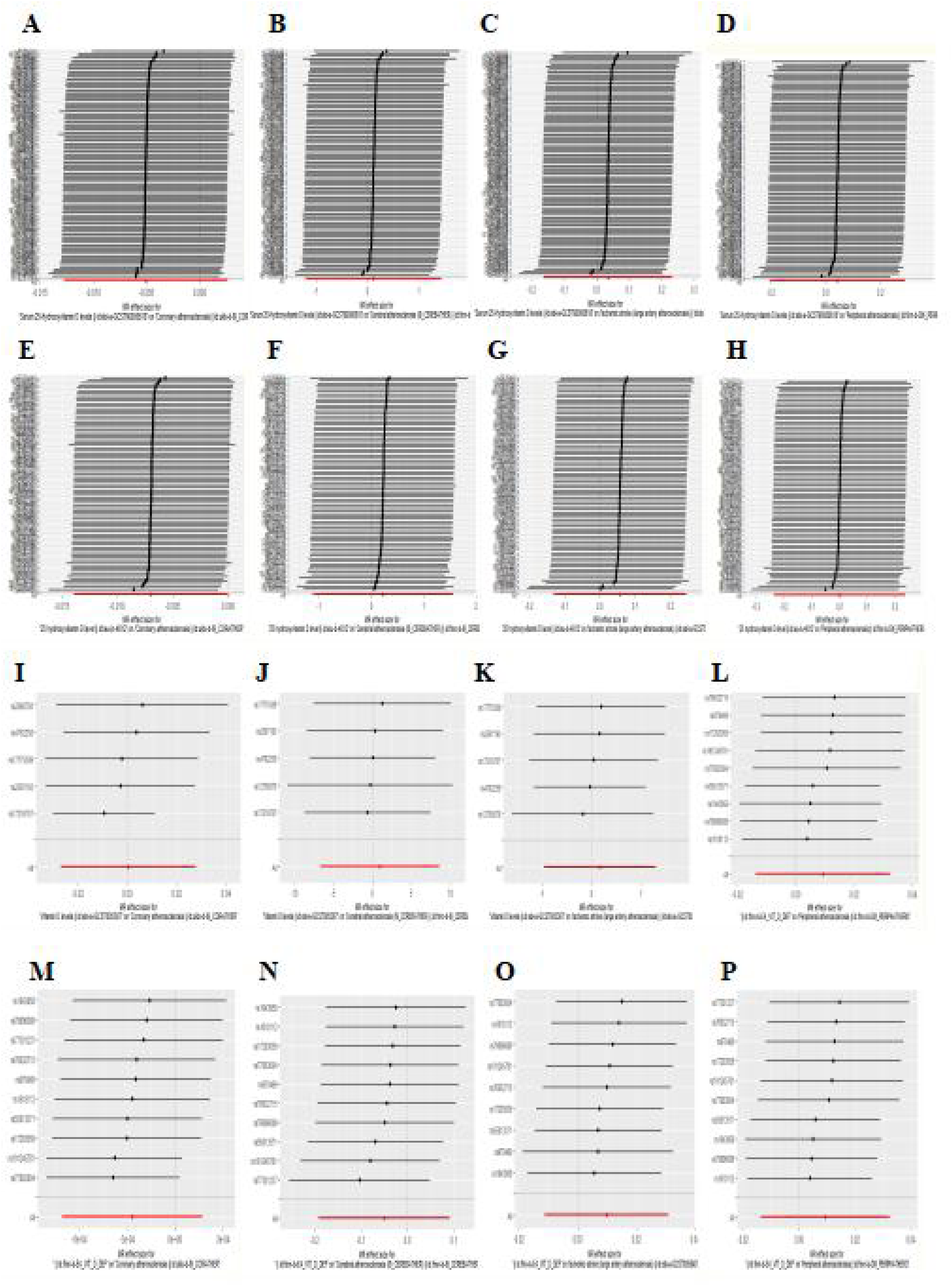
Vitamin D levels and atherosclerosis MR analysis leave-one-out plots ABCD: ID ebi-a_GCST90000618: A. Coronary atherosclerosis, B. Cerebral atherosclerosis, C. Large artery atherosclerosis, D. Peripheral atherosclerosis; EFGH: ID ieu_b_4812: E. Coronary atherosclerosis, F. Cerebral atherosclerosis, G. Large artery atherosclerosis, H. Peripheral artery atherosclerosis; IJKL: ID ebi-a-GCST005367: I. Coronary artery atherosclerosis, J. Cerebral artery atherosclerosis, K. Large artery atherosclerosis, L. Peripheral artery atherosclerosis; MNOP: ID finn-b-E4_VIT_D_DEF: M. Coronary artery atherosclerosis, N. Cerebral artery atherosclerosis sclerosis, O. aortic atherosclerosis, P. peripheral atherosclerosis.

**Table 1.**
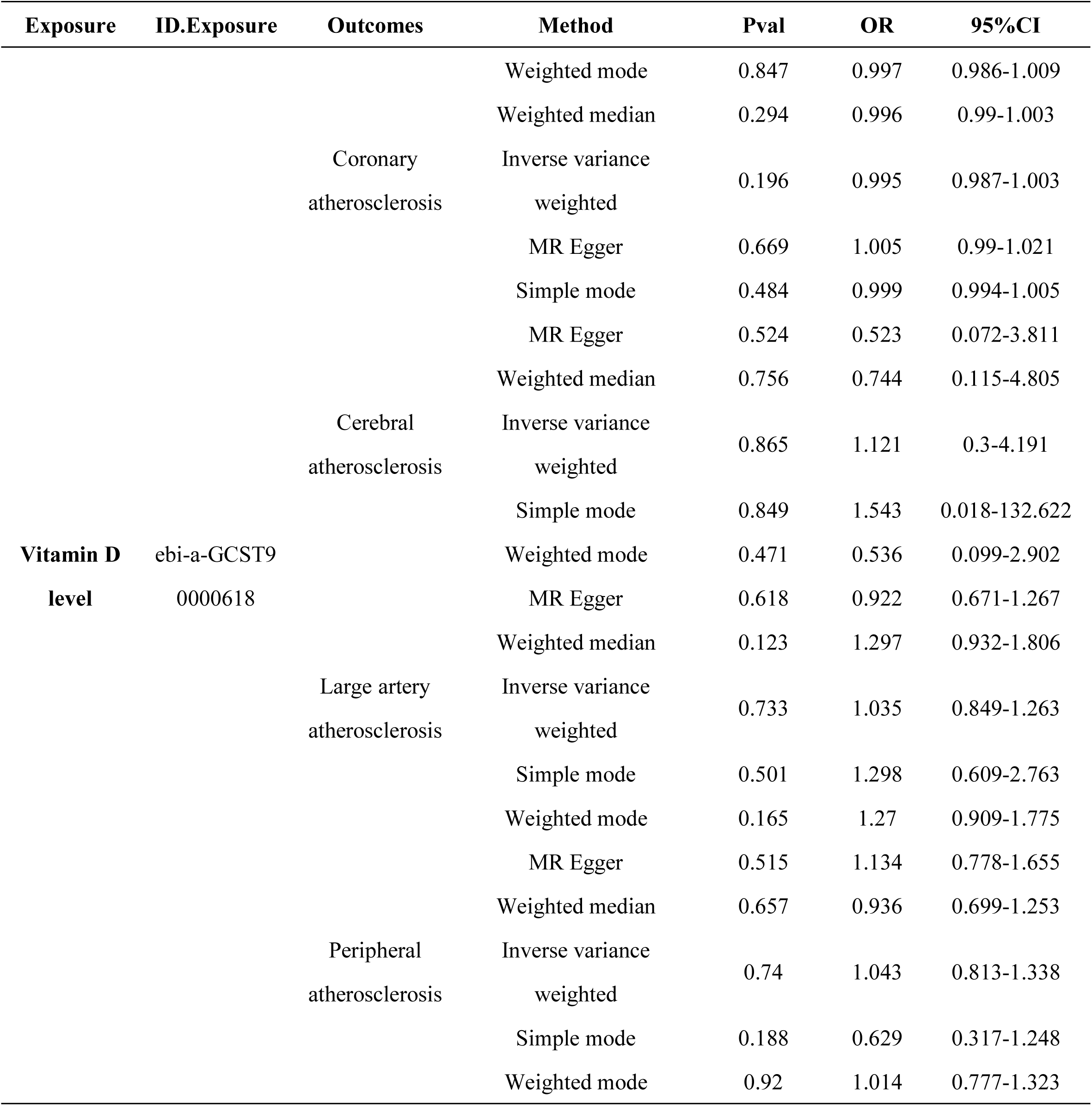
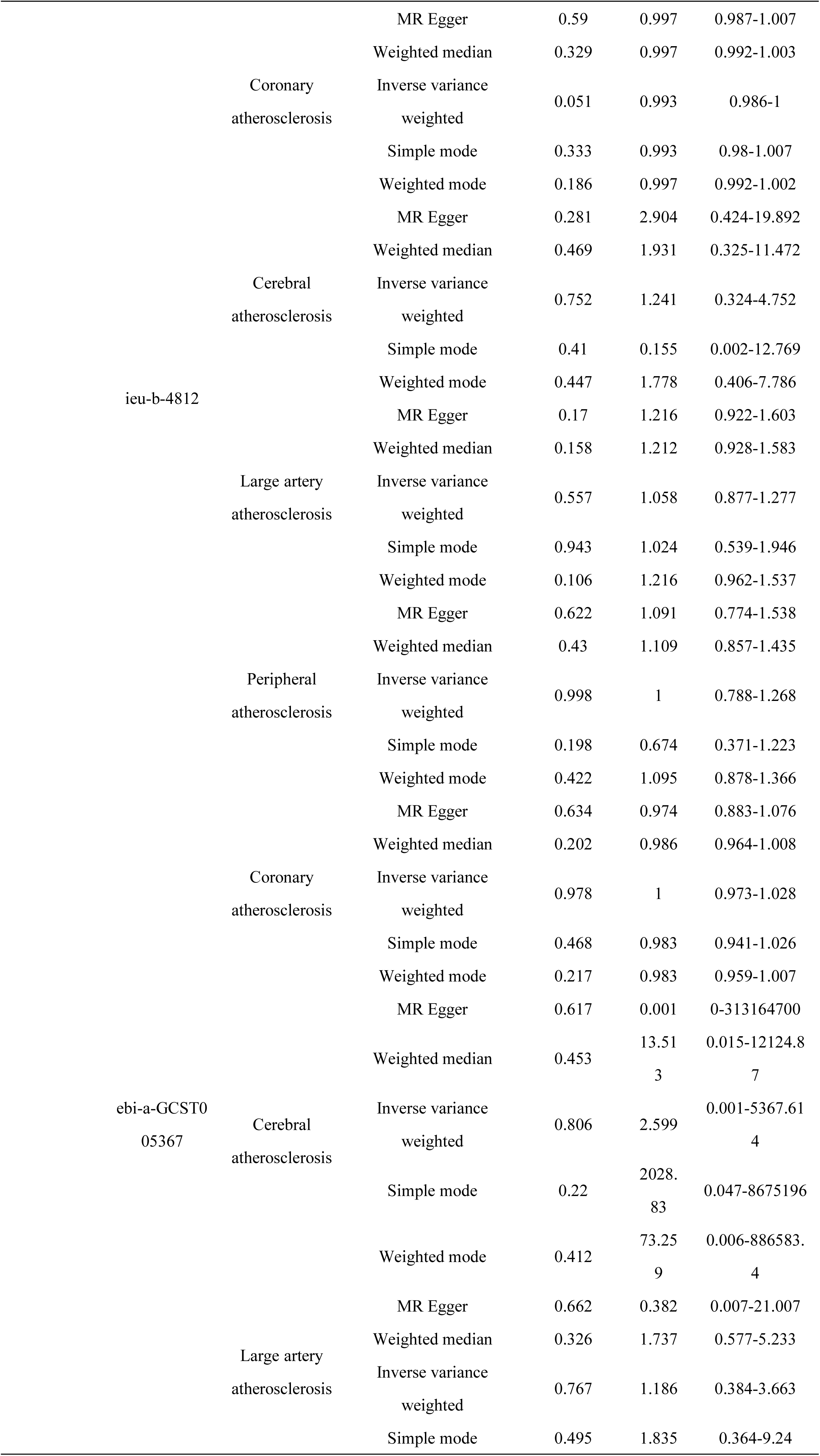
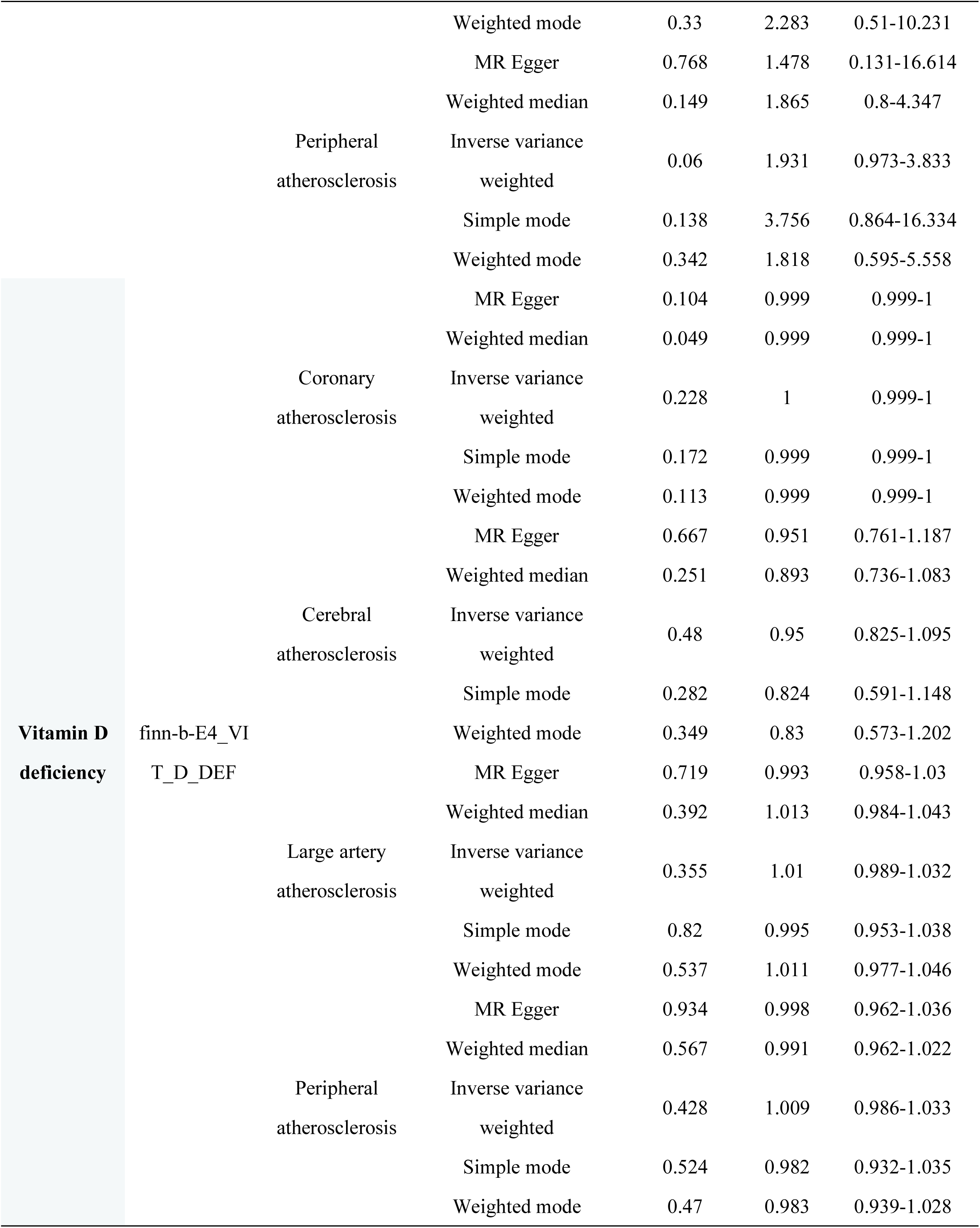
MR analysis results.

### 3.3 Sensitivity analysis

Analyzed by Cochran’s Q test we found that there was heterogeneity of SNPs in individual sample data (P < 0.05), as shown in Table 2. To make the data showing heterogeneity more obvious, we marked them in yellow in the table. This indicates that there was bias between the exposure factors in the study and the outcome sample data, which affected the results of the analyses and made it difficult to form a reliable causal association. We excluded the SNPs with high outliers from the leave-one-out analysis, and the results still suggested that there was evidence of heterogeneity. MR-Egger intercept analysis, the results showed that there was no evidence of horizontal multivariate validity existing in the sample data, i.e., P > 0.05, suggesting that there was a strong correlation between the SNPs of the sample data selected for the present study and the exposures, which was consistent with the MR analysis that the instrumental variables acted on the outcomes only through the exposure factors. Hypothesis. To make the results more reliable, we followed up with MR-PRESSO analyses, and no evidence suggestive of horizontal pleiotropy was found in the results either.

**Table 2.**
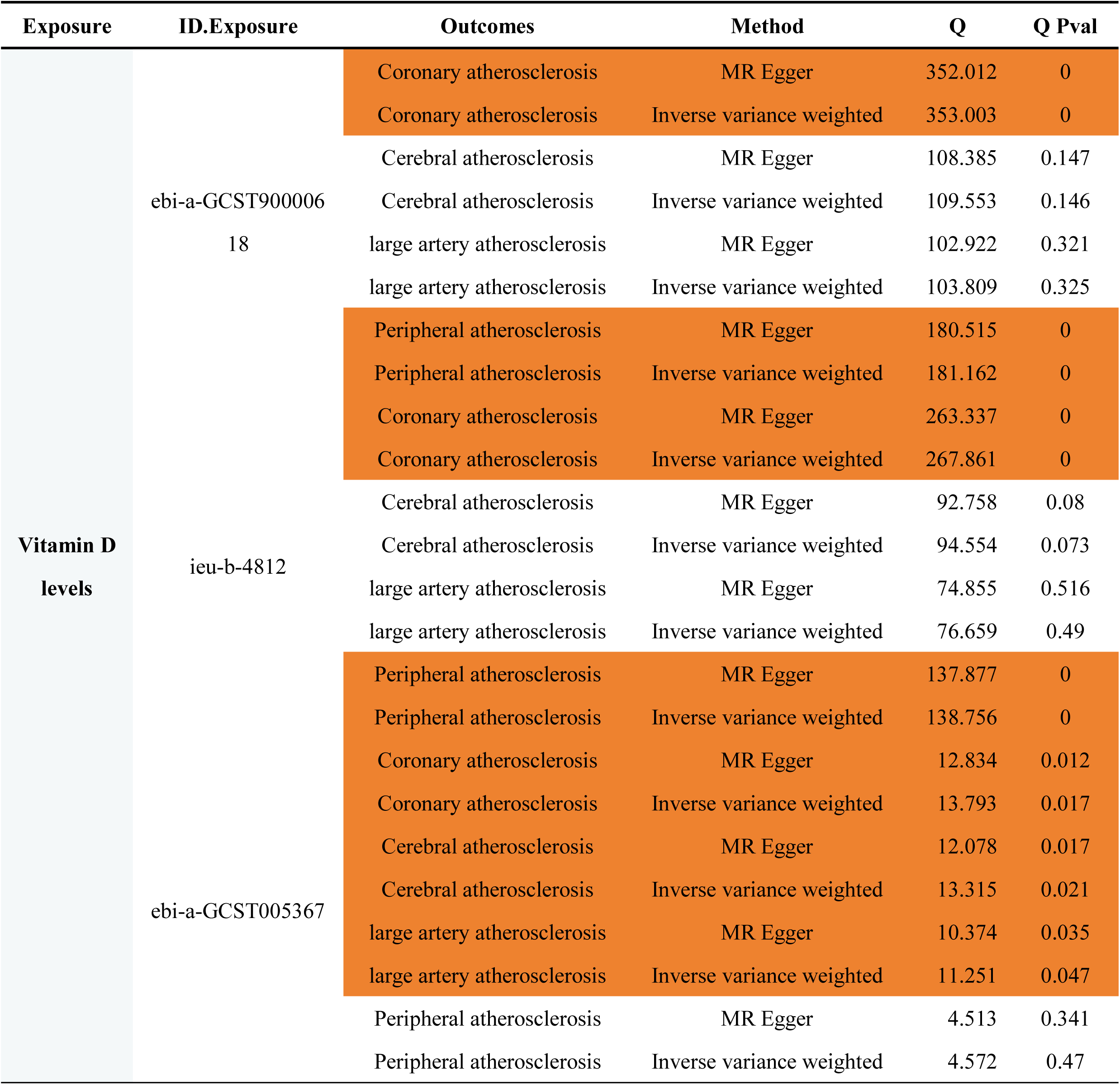
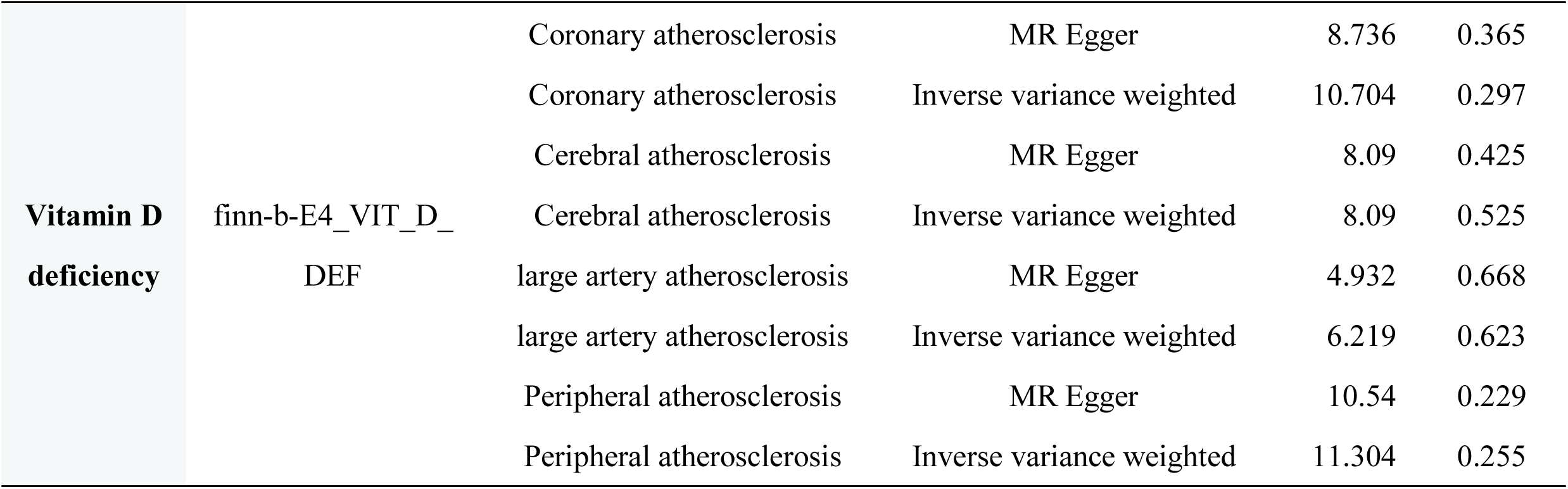
Results of Cochran’s Q test.

### 3.4 Reverse MR analyses

To assess whether there was an inverse causal association between vitamin D levels, vitamin D deficiency, and the four atherosclerosis, we performed separate MR analyses with the original exposure factors (three vitamin D levels, vitamin D deficiency) as the outcome and the original outcome (four atherosclerosis) as the exposure factors. The results showed an inverse causal association between peripheral atherosclerosis and vitamin D deficiency (IVW P=0.042), whereas no inverse causal association existed between the other three types of atherosclerosis and vitamin D levels, and vitamin D deficiency (IVW P>0.05). That is, peripheral atherosclerosis can trigger vitamin D deficiency, with a correlation of OR=1.898, 95% CI=1.024-3.518. Sensitivity analyses resulted in no evidence of heterogeneity among the corresponding SNPs (Q_Pval=0.910), as shown in **Table 3**. Horizontal pleiotropy was assessed by the MR-Egger intercept method, and again no evidence of the existence of horizontal pleiotropy was found (P=0.765), meaning that the corresponding SNPs were reasonably selected and highly associated with the exposure factors. The leave-one-out method of analysis showed that the corresponding SNPs were robust (all located to the right of 0), as shown in **Figure 4** and **Figure 5**.

**Table 3.** Positive results of inverse MR analysis.

| Exposure | Outcome | Method | Pval | OR (95%CI) |
| --- | --- | --- | --- | --- |
| Peripheral atherosclerosis | Vitamin D deficiency | MR Egger | 0.765 | 1.641(0.078-34.322) |
|  |  | Weighted median | 0.066 | 2.022(0.953-4.288) |
|  |  | Inverse variance weighted | 0.041 | 1.898(1.024-3.518) |
|  |  | Simple mode | 0.2 | 2.383(0.753-7.545) |
|  |  | Weighted mode | 0.408 | 1.599(0.578-4.425) |

**Figure 5.**
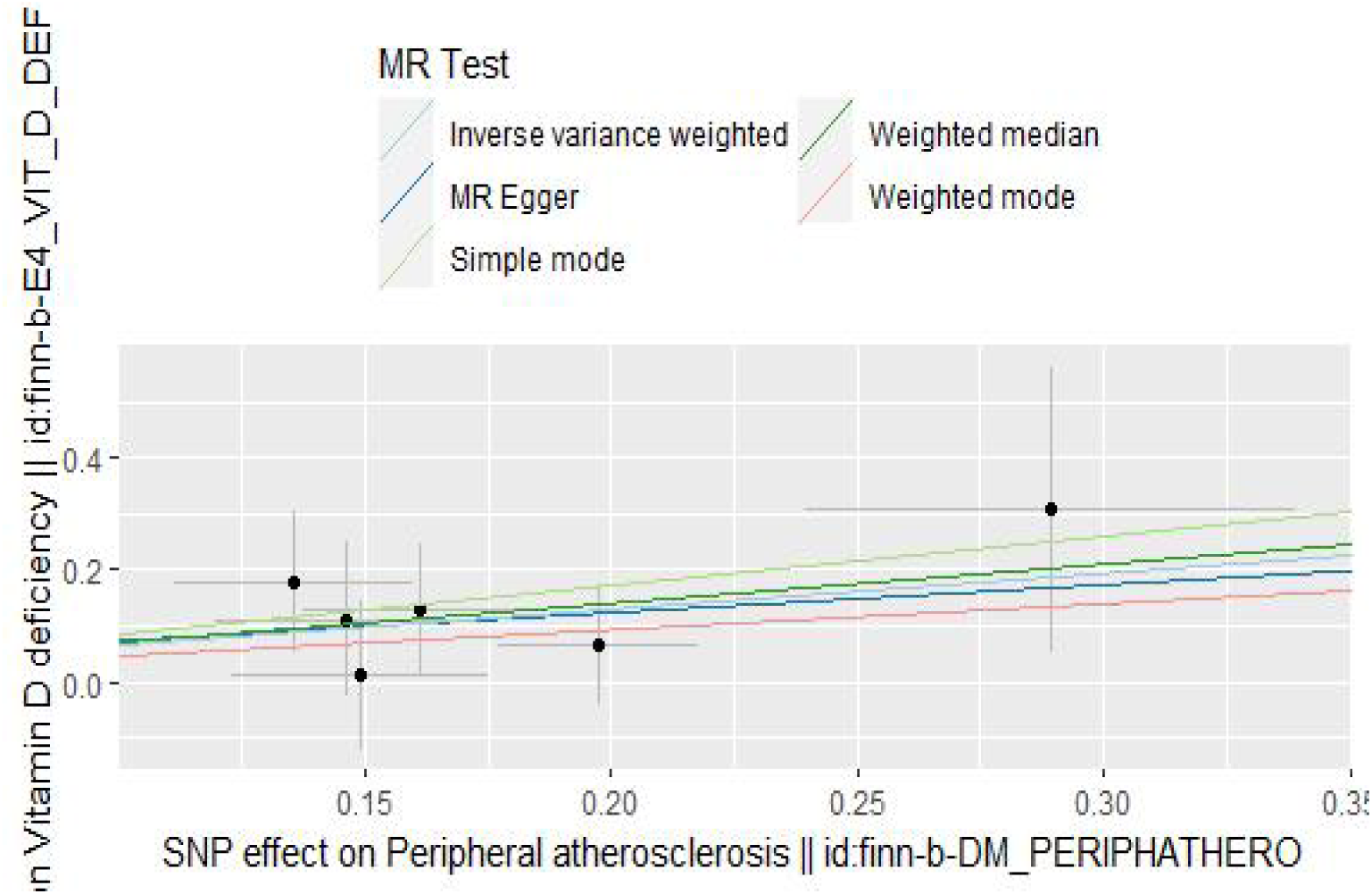
Scatter plot of the relationship between peripheral atherosclerosis and vitamin D deficiency.

**Figure 6.**
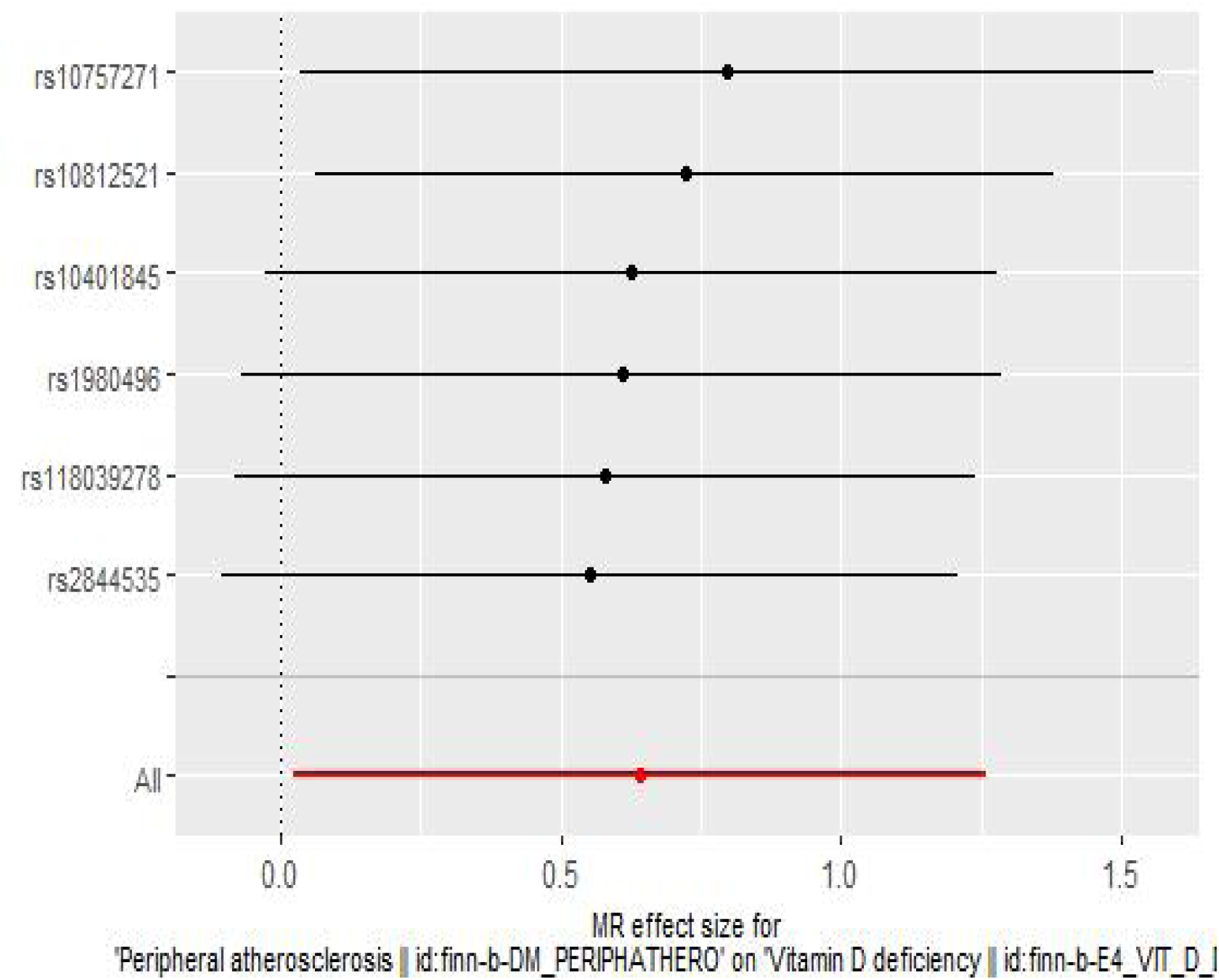
Leave-one-out plot of the relationship between peripheral atherosclerosis and vitamin D deficiency.

## 4 Discussion

In this study, we applied a two-sample bidirectional MR analysis to explore the causal link between vitamin D and AS. Our findings suggest that there is no positive causal relationship between vitamin D level status and AS. However, we unexpectedly found a reverse causal relationship between vitamin deficiency and peripheral atherosclerosis. This finding suggests that the development of peripheral atherosclerosis can lead to vitamin level deficiency, i.e., peripheral atherosclerosis increases the prevalence of vitamin D deficiency. Otherwise, there was no evidence of reverse causality between vitamin D status and its three atherosclerosis.

The progression of AS involves the interaction of inflammatory mediators, and immune molecules, and the role played by vitamins in inflammation and immunity is well-established and clear [12]. The results of the analysis showed heterogeneity among SNPs, and the heterogeneity was still present after excluding the SNAs with significant bias, suggesting that a single SNP had a large impact on the results of the study, and it is possible that vitamin D does not have a direct effect on AS, but rather acts through mechanisms involved in inflammation, and immune response. It is also possible that the GWAS data on vitamin D status do not distinguish between the circulating form (25(OH)D) and the active form 1,25(OH)2D, which needs to be converted to the active form by the enzyme 1-alpha-hydroxylase in the kidneys to act. In addition to this, it has been found that the causality between vitamin D deficiency and cardiovascular disease is non-linear, i.e., it does not produce an effect for a single factor [13].AS, as a major contributing factor to cardiovascular disease, may have this associative property with vitamin D. The association was not found to be significant in the absence of vitamin D deficiency. The inverse MR analysis did not show heterogeneity, and the corresponding leave-one-out results showed no bias, suggesting that the data were stable and the conclusions were reliable.

Vitamin D deficiency is often considered to be associated with diseases such as osteoporosis and fractures, and recent evidence confirms that there is also an association between vitamin D deficiency and deaths due to cardiovascular disease [14, 15]. In addition, there is evidence that vitamin D deficiency is associated with risk factors for cardiovascular disease, such as smoking, alcohol consumption, and hypertension [16–21]. Vitamin D also plays a role in protection against cardiovascular disease, such as anti-inflammatory, inhibition of plaque formation, and protection of the vascular endothelium [22–24]. Michal L. Melamed et al. conducted a cross-sectional study on the relationship between vitamin D levels and all-cause mortality in 13,331 individuals from the NHANES III survey in the United States, and showed that smoking, body mass index (BMI), diabetes, and other factors contribute to the risk of cardiovascular disease [21]. BMI, and diabetes were all independently associated with vitamin D deficiency, and among cardiovascular diseases, deaths from AS were as high as 76%, multivariate regression analyses were used after adjusting the data, which showed that cardiovascular mortality due to vitamin D deficiency was still high [25]. Related studies have shown that vitamin D deficiency is associated with coronary artery disease (CAD) due to AS. Yashwant Agrawal et al. performed a regression analysis based on NIS data from 2008 to 2012 in the United States and found that patients with VDD were approximately 1.13 times more likely to develop CAD compared to those without VDD, and the mortality rate for patients with CAD due to VDD was almost 1.81 times more likely to have VDD than those without VDD.

Lipid levels are a major factor affecting AS, Jorde, R et al. comprehensively explored the relationship between vitamin D and blood quality, and relevant cross-sectional, ecological studies were sufficient to show that vitamin D can lead to lower lipid levels [26]. In animal experiments by Peter F Schnatz et al, arterial plaque formation was more pronounced in monkeys with low levels of vitamin D receptor than in monkeys with high levels, and there was a correlation between the thickness size of the plaque and the vitamin D receptor [27]. Palanikumar Gunasekar et al, gave different levels of vitamin D to pigs, and those with high levels of vitamin D showed a significant decrease in inflammatory M1 macrophages significantly reduced in pigs with high levels of vitamin D, while anti-inflammatory M2 macrophages showed a corresponding increase, and M1 macrophages were significantly increased in vitamin D-deficient pigs, which suggests that vitamin D is positively correlated with the suppression of anti-inflammatory M2 macrophages in AS and reduces the level of inflammatory M1 macrophages [28]. Suggesting that the effect of vitamin D on AS is likely to act by affecting macrophages. However, our present forward MR analysis did not provide evidence that vitamin D can lead to an increased risk of atherosclerosis.

Our reverse MR analysis showed that peripheral atherosclerosis can lead to an increased risk of developing vitamin D deficiency. The subsequent sensitivity and leave-one-out analyses did not reveal any data bias, suggesting that our results have a certain degree of confidence. In this regard, some studies have confirmed the association between peripheral atherosclerosis and osteoporosis-causing factor osteoprotegerin kinase (OPG), and OPG levels are significantly elevated in patients with peripheral atherosclerosis [29]. It has also been shown that there is a positive association between some vascular calcification markers and peripheral arterial disease, but there is still controversy among scholars about the accuracy and credibility of the results [30]. However, individuals with the peripheral arterial disease generally have low levels of vitamin D compared to those without peripheral arterial disease, as demonstrated in studies by Veronese N et al. and Rapson IR et al [31, 32]. Thus, our findings further suggest a causal relationship between the two and thus help in clinical follow-up for targeted research and treatment.

## 5 Limitations of this study

In this MR study, the data of the GWAS samples we selected were from European groups, and our results may not be able to be extrapolated to other ethnic populations; after all, different ethnic genes may lead to different analysis results. Secondly, in the course of our study, there is a bias in the SNP data, which may have an impact on the accuracy of the analyses and the reliability of the results.

## 6 Conclusion

Although scholars’ studies have suggested an association between vitamin D and AS, our MR analyses did not suggest evidence of a causal relationship. It is worth noting that we found peripheral atherosclerosis to be the cause of vitamin D deficiency in our inverse MR, which undoubtedly provides a basis and reference for future research and clinical treatment protocols for vitamin D deficiency.

## Supplementary Material

The following supporting information can be seen at TableS1:Supplementary Table 1; TableS2: checklist-fillable.

## Author contributions

Original writing, Liu.Chunhui. and Huang.Xvpeng.; Supervison, Jin.Hongguang. and Huang.Yongsheng.

## Institutional Review Board Statement

The data of the cases included in this study have obtained authorized consent from the patients themselves and the approval of the ethical review body, and there is no need for further ethical review.

## Informed Consent Statement

Informed consent was obtained from all subjects involved in the study.

## Data Availability Statement

The data used in this article were obtained from the GWAS public database, which is available online at https://gwas.mrcieu.ac.uk/.

